# Genomic Landscape of Early-Onset and Familial Latin American Parkinson’s Patients

**DOI:** 10.64898/2026.09.16.26359514

**Authors:** Emily Waldo, Henry Mauricio Chaparro-Solano, Mariam Isayan, Thiago Peixoto Leal, Felipe Duarte-Zambrano, Miguel Inca-Martinez, Janvi Ramchandra, Maria Rivera Paz, Mary B. Makarious, Carlos Hernandez, Emilia M Gatto, Natalia Gonzalez Rojas, Martin Emiliano Cesarini, Bruno Lopes Santos-Lobato, Grace Helena Letro, Jorge Luis Orozco, Beatriz Munoz Ospina, Pedro Chana-Cuevas, Natalia Andrea Rojas, David Aguillon, Valentina Muller, Pedro Braga-Neto, Mayela Rodríguez-Violante, Amin Cervantes-Arriaga, Artur F S Schuh, Mario Cornejo-Olivas, Koni Mejia Rojas, Cintia Armas, Ángel Viñuela, Alan Osvaldo Espinal Martinez, Vitor Tumas, Vanderci Borges, Cesar Luis Avila, Patricio Olguin, Sarael Alcauter, Marcelo Kauffman, Dolores Gonzalez-Moron, Susana Lissette Peña, Ignacio F. Mata, the Latin American Research Consortium on the Genetics of Parkinson’s Disease

## Abstract

**Background:** Parkinson’s disease (PD), the most common neurodegenerative movement disorder, is commonly thought of as an aging and sporadic disease; however, 5-14% of individuals experience disease onset before the age of 50 years (early-onset PD; EOPD) and about 20% have a positive family history. In the case of both EOPD and people with a family history of PD, evidence suggests higher rates of a disease-causing genetic contribution.

**Objectives:** Our goal was to perform a variant screening of seven PD-related genes and 21 genes associated with related parkinsonian disorders to identify pathogenic/likely pathogenic variants and variants of uncertain significance within EOPD and family-history-positive individuals within the Latin American Research Consortium on the Genetics of PD (LARGE-PD).

**Methods:** We performed short-read whole-genome sequencing on a subset of 263 individuals with EOPD and/or a familial history of PD who had no known pathogenic PD variant in genotyping data.

**Results:** Of the analyzed individuals, a monogenic burden in primary and secondary PD genes due to pathogenic or likely pathogenic variants for PD was observed in 4.7%, an additional 5.5% had pathogenic or likely pathogenic variants for Gaucher’s disease, and 2.7% had known risk variants in *GBA1*.

**Conclusions:** By expanding the reported spectrum of disease-associated variants in a Latin American population, we identified previously unreported or underrepresented variants that would likely be missed by array-based or targeted screening approaches and contribute to the characterization of EOPD and familial PD in Latin America.

## INTRODUCTION

Parkinson’s disease (PD) is a neurodegenerative movement disorder that most commonly presents after 60 years of age, with incidence increasing with age (1). Approximately 5-14% of people living with PD (PwP) experience disease onset before the age of 50, referred to as early-onset PD (EOPD) (2). Although both EOPD and late-onset PD (LOPD) arise from complex interactions between genetic, environmental, and lifestyle factors, pathogenic or likely pathogenic variants in PD-associated genes account for a substantially greater proportion of EOPD than LOPD. The estimated monogenic diagnostic yields are approximately 18.4% and 11.9%, respectively (3). Monogenic PD is defined as PD where the genetic contribution of one or a set of variants in one gene is disease-causing. Additionally, around 20% of PwP report a positive family history of PD in a 1st or 2nd degree relative (4), a subset that overlaps substantially with EOPD, with 20% of individuals with EOPD reporting a positive family history of PD as well (5). Consequently, current clinical guidelines recommend prioritizing genetic testing in individuals with EOPD and those with a positive family history of PD (6).

The age-standardized incidence and prevalence of EOPD vary by region and population, with the highest rates in Andean Latin America and East Asia, with age-standardized incidence rates of 5.10 and 4.54 per 100,000 people, respectively (7). When broken down further, three of the top four EOPD age-standardized incidence rates by country were from Latin America (per 100,000 people; Bolivia: 5.1, Ecuador: 4.9, and Peru: 5.2) (7). The factors underlying these geographic differences remain poorly understood but may reflect differences in genetic architecture, environmental exposures, lifestyle, or a combination of these factors. Understanding the contribution of each of these components requires comprehensive genetic characterization of populations with the highest EOPD disease burden. However, most PD genetics research has been conducted in individuals of European and Asian ancestry, limiting the comprehensive understanding of the genetic architecture of PD in underrepresented populations, including Latin Americans (8,9). This gap is particularly significant for the monogenic landscape of EOPD. Although researchers have investigated familial PD in Latin America (8,9), the prevalence and spectrum of pathogenic variants causing PD and related parkinsonian disorders remain incompletely characterized.

The impact of this incomplete characterization is that current PD clinical genetic testing strategies frequently rely on targeted sequencing panels, in which pathogenic variants are primarily classified based on (10). These approaches may fail to capture population-specific pathogenic variants, which may be classified as variants of uncertain significance (VUS), reducing their diagnostic yield in underrepresented populations such as Latin Americans. Similarly, though some genotyping arrays, including the NeuroBooster Array (NBA), incorporate a limited number of established pathogenic variants alongside approximately 1.9 million genome-wide markers and ∼95,000 neurological disease-focused variants (11), their coverage of rare disease-causing variants, as well as complex genetic regions like the *GBA1* region, which includes a pseudogene, remains incomplete. Therefore, the absence of a pathogenic variant on targeted panels or genotyping arrays cannot exclude a monogenic etiology, particularly in genetically understudied populations.

Whole-genome sequencing (WGS) provides an unbiased approach for identifying both known and novel pathogenic variants across established PD and parkinsonism-associated genes. Defining the monogenic landscape of EOPD and familial PD in Latin American populations is essential for improving molecular diagnosis, identifying individuals who may benefit from gene-targeted therapeutic trials, refining estimates of the contribution of monogenic disease, and informing genetic counseling and prognosis.

To address this gap, we conducted a comprehensive genetic variant screening using short-read WGS data from a cohort of over 250 Latin American individuals with EOPD and/or family history of PD. All included individuals previously tested negative for a known pathogenic or likely pathogenic (P/LP) variant in a primary PD gene (*LRRK2*, *PARK7*, *PINK1*, *PRKN*, *SNCA*, or *VPS35*) using the NBA. This screen re-evaluated the primary PD genes for additional disease-relevant variants and extended coverage to a broader panel of secondary genes implicated in other parkinsonisms. We present the monogenic burden attributable to P/LP variants and evaluate how this burden changes when VUS are also considered.

## METHODS

### Cohort and Participant Inclusion Criteria

The Latin American Research consortium on the Genetics of PD (LARGE-PD) is a multinational consortium spanning 52 active sites across 15 countries in the Americas and Caribbean that collects genetic, environmental, and lifestyle data on PwP and unaffected controls (12). Inclusion criteria for this study included: participants were 1) at least 18 years old at recruitment, 2) reported themselves or a 1st or 2nd degree relative of Hispanic/Latin American origin, and 3) diagnosed with PD based on UK Brain Bank and/or International Parkinson Movement Disorders Society (IPMDS) diagnostic criteria. We have described LARGE-PD in detail previously (12,13).

All participants provided written informed consent. This study was approved by the Institutional Review Board (IRB) of the Cleveland Clinic Foundation (IRB #19-340), and ethical approval was obtained from the local IRB of all participating LARGE-PD sites.

### Participant Filtering

DNA was extracted from blood (n=1,222) or buccal swab (n=86) samples collected between 2019 and 2024 and genotyped with the NBA from 1,308 individuals falling into at least one of two clinically high-risk subgroups for genetic causes: 1) age at onset of motor symptoms ≤50 years (EOPD; n=742) or 2) PwP who also had a history of at least one 1st or 2nd degree relative diagnosed with PD (FHx+, n=349), 3) or have both EOPD and FHx+ (n=217).

To filter out the most common and well-established monogenic PD variants to maximize the likelihood of observing population-specific or unreported variants, a list of previously reported P/LP variants in primary PD genes (*LRRK2*, *PARK7*, *PINK1*, *PRKN*, *SNCA*, and *VPS35*, Supplementary Table 1) was extracted from the NBA data, and carriers were excluded for the next step. Though variants in *GBA1* are associated with PD, they are primarily classified as risk alleles, rather than disease-causing, so individuals were not excluded based on *GBA1* variant carrier status, as there may be co-occurrence of other causative monogenic variants (14). Variants in *GBA1* are also sometimes poorly captured on genotyping arrays; therefore, we aimed to validate those found in genotyping data via sequencing. From the 1,272 individuals remaining after filtering, a pilot subset of 263 individuals (biospecimen from blood: 259, biospecimen from buccal swab: 4) was sequenced with Illumina NovaSeq as part of an initial sequencing phase (Illumina: NovaSeq 6000 Sequencing System (RRID: SCR_016387)). Individuals with mean coverage >30x were retained (Figure 1).

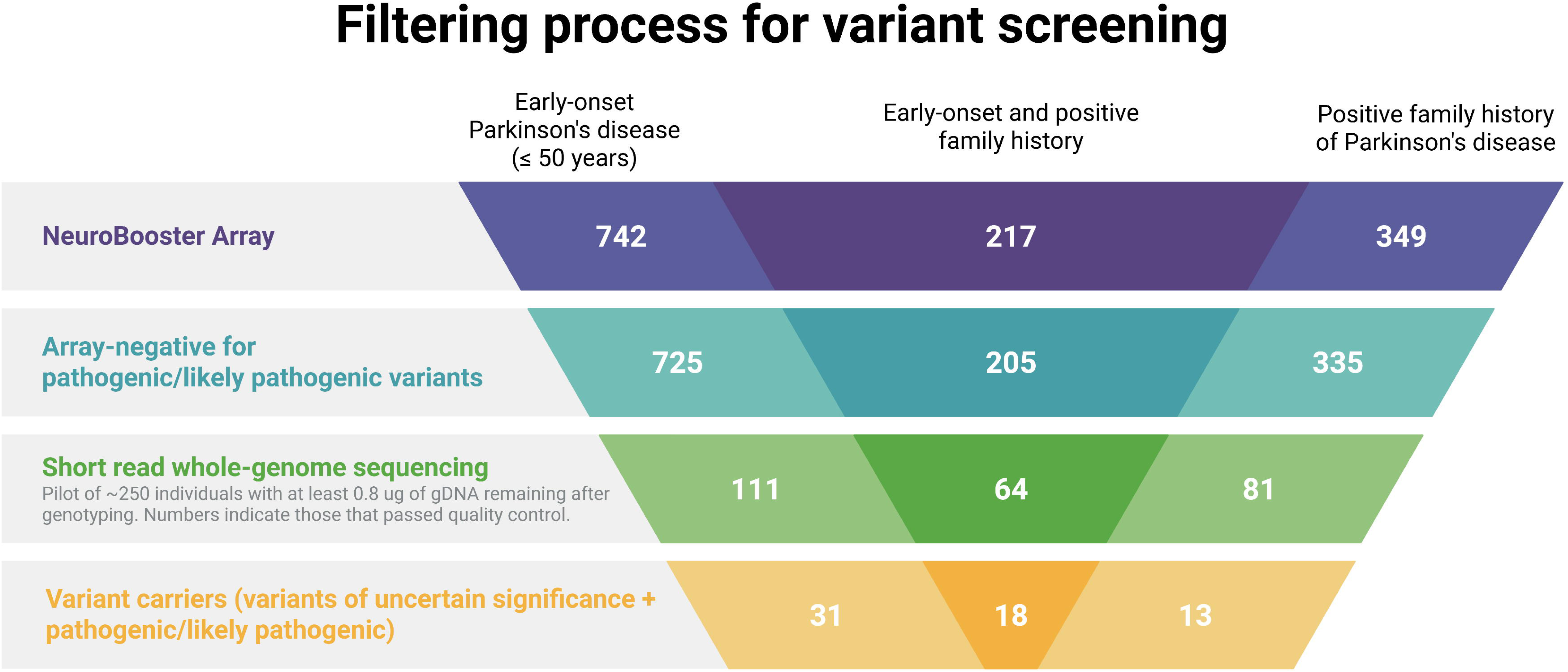

### Variant Calling and Annotation

Variant calling, normalization, and allele alignment of single-nucleotide variants (SNVs), insertion-deletions (indels), and copy-number variants (CNVs) were performed with Illumina DRAGEN Germline v.4.4.6, and individuals were joint called with DRAGEN PopGen v.4.3.6 (15) in accordance with GRCh38.

Variants were annotated for functional consequence, *in silico* deleteriousness, population frequency, and known clinical significance using a standard multi-tool pipeline (VEP (16), snpEff (17), ANNOVAR (18), dbNSFP (19), CADD (20), gnomAD (21), ClinVar (22); full details in Supplementary Methods).

### Variant Prioritization

For the EOPD and/or FHx+ individuals in which no monogenic cause was identified after filtering with the NBA, WGS variant calls were screened in the seven primary PD genes (the six above plus *GBA1*), plus 21 secondary genes associated with other parkinsonian syndromes, for P/LP and VUS (23)(Supplementary Table 2). *GBA1* was not used for array-based exclusion but was included in the WGS screen due to prior evidence of its impact as a modifier, especially for *LRRK2* variants. Genomic regions were extracted from the Entrez Gene database (24).

Variant inclusion criteria were defined according to the mode of inheritance (MOI) of the associated gene and phenotype. Heterozygous variants in autosomal dominant (AD) genes were eligible for inclusion, whereas variants in autosomal recessive (AR) genes were considered if they were in a homozygous or compound heterozygous state. For compound heterozygous variants, a second SNV, indel, or CNV was required within the same gene in *trans* orientation, as determined by statistical phasing, with a classification of VUS or higher under American College of Medical Genetics and Genomics and the Association for Molecular Pathology (ACMG/AMP) criteria (25). For SNVs and indels, compound heterozygotes were phased with SHAPEIT5 phase_common to evaluate variant orientation (26).

Sequencing metrics further filtered variants. To ensure only high-quality variants are reported, per-sample genotype (GT), depth (DP), and genotype quality (GQ) metrics from the joint-called VCF file were extracted. Carriers with GQ ≥ 30 and DP ≥ 20 were considered to pass the quality thresholds, except when an SNV and CNV were located within the same exon, in which case DP metrics were confirmed to be consistent with single-copy coverage. In Supplementary Table 3, average GQ and DP metrics for variants of interest are reported.

The final prioritization was based on classification of variant impact. The list of prioritized variants was created based on the following criteria: (1) known pathogenic variants reported in ClinVar, (2) variants classified as VUS or higher in Varsome (https://varsome.com/) and Franklin (https://franklin.genoox.com/) databases, (3) VUS with CADD score > 15 and allele frequency reported in gnomAD < 1%. As an exception, we selected variants in the *GBA1* gene from the GBA1-PD browser regardless of reported frequency, based on a CADD score > 15. Retention of those variants was based on the frequency of pathogenic variants in the *GBA1* gene in the heterozygous state, which can vary across populations, ranging from up to 1% in the general population to 8% in Ashkenazi Jewish ancestry (14).

The CNVs affecting one or more exons of *PARK7*, *PINK1*, *PRKN*, and *SNCA* were retained for interpretation. CNV calls were filtered using DRAGEN’s default filtering criteria.

### Variant Classification and Interpretation

The SNVs remaining after filtering criteria were applied and consistent with the MOI of the associated phenotype were classified according to the ACMG/AMP guidelines for the interpretation of sequence variants (25). The MDSGene database, developed by the IPMDS (27), was used as a disease-specific resource to determine whether candidate variants had been previously reported in individuals with PD or other forms of parkinsonism, the populations in which they had been identified, and whether functional evidence supported their classification.

The primary analysis focused on variants classified as P/LP, while VUS were also evaluated and reported separately.

CNVs were evaluated in the context of the associated inheritance pattern, available evidence from ClinGen and DECIPHER (28), and ACMG/ClinGen recommendations for CNV interpretation (29). Particular attention was given to those CNVs that were in combination with another P/LP variant in the same gene when consistent with a recessive MOI.

### Presence in the Global Parkinson’s Genetics Program

We assessed the presence of variants of interest identified in LARGE-PD across the Global Parkinson’s Genetics Program (GP2) through the publicly available GP2 Browser (https://gp2.broadinstitute.org/). The GP2 is an initiative spanning over 415 cohorts worldwide, collecting historically underrepresented populations (30). The latest data release (GP2, Release 12, 10.5281/zenodo.20932193) features over 60k short-read WGS data from individuals with PD and healthy controls. We screened the variants of interest identified in our study in the GP2 Browser. We exported variants in genes of interest, then filtered and reported the frequencies in PD cases and controls for variants present in the GP2 dataset. Other phenotypes were not included in our validation (e.g., PSP, DLB, MSA).

## RESULTS

### Cohort description

A total of 263 individuals from LARGE-PD from eight Latin American countries (Argentina, Brazil, Chile, Colombia, El Salvador, Mexico, Peru, and the United States (Puerto Rico)) were whole-genome sequenced. After filtering by sequencing coverage, 256 participants were retained for analysis. Of these, 111 (43.4%) had EOPD, 81 (31.6%) had FHx+, and 64 (25.0%) had both EOPD and FHx+. Group-specific demographic characteristics are presented in Table 1.

**Table 1.** Demographic table of sequenced individuals by country and analysis group.

|  |  |  |  |  |  |  |  |  |  |
| --- | --- | --- | --- | --- | --- | --- | --- | --- | --- |
| **Country** | **EOPD** |  |  | **Family history positive** |  |  | **EOPD and family history** |  |  |
|  | **Count** | **Average Age at Onset** | **Percent Males** | **Count** | **Average Age at Onset** | **Percent Males** | **Count** | **Average Age at Onset** | **Percent Males** |
| Argentina | 25 | 34 | 52.00% | 16 | 58 | 37.50% | 19 | 42.1 | 42.10% |
| Brazil | 16 | 36.5 | 68.80% | 1 | 57 | 100.00% | 4 | 35.3 | 100.00% |
| Chile | 11 | 37.2 | 72.70% | 4 | 59.5 | 100.00% | 6 | 34.3 | 66.70% |
| Colombia | 17 | 32.7 | 76.50% | 25 | 66.4 | 76.00% | 13 | 38 | 53.80% |
| El Salvador | 1 | 38 | 100.00% | 0 |  |  | 0 |  |  |
| Mexico | 13 | 34.8 | 69.20% | 0 |  |  | 7 | 32.1 | 28.60% |
| Peru | 23 | 32 | 30.40% | 22 | 64.7 | 40.90% | 11 | 37.5 | 36.40% |
| United States (Puerto Rico) | 5 | 38.6 | 40.00% | 13 | 59.8 | 84.60% | 4 | 42.8 | 50.00% |
| **TOTAL** | **111** | **34.4** | **57.70%** | **81** | **62.8** | **61.70%** | **64** | **38.3** | **48.40%** |
| EOPD: early-onset Parkinson's disease |  |  |  |  |  |  |  |  |  |

### Yield

From 256 individuals of NBA-negative EOPD and FHx+ individuals, a monogenic burden in primary and secondary PD genes due to P/LP variants is shown in 4.7% (EOPD: 4.4%; FHx+: 3.7%; EOPD and FHx+: 4.7%) of PwP and an additional 5.5% (EOPD: 1.8%; FHx+: 0%; EOPD and FHx+: 1.6%) of variants P/LP for Gaucher’s disease in *GBA1* and 2.7% of individuals with known *GBA1* risk variants (EOPD: 0.01%, FHx+: 2.5%, EOPD and FHx+: 6.3%).

### Pathogenic and Likely Pathogenic variants

Of the 17 unique disease-causing P/LP SNVs, thirteen variants were identified in established PD-associated genes (*GBA1*, *PRKN*, *LRRK2*, and *VPS13C*), whereas four were found in genes associated with other forms of parkinsonism (*MAPT* and *POLG*). The identified variants included missense, nonsense, frameshift, splice-site, and synonymous splice-altering variants, illustrating the molecular heterogeneity of monogenic PD in this cohort.

Among the PD-associated genes, the most frequently observed P/LP variants were identified in *GBA1*, though it should be noted that these variants are P/LP for Gaucher’s disease not PD.

The two most common variants were c.1448T>C (p.Leu483Pro) and c.1226A>G (p.Asn409Ser), each detected in five heterozygous participants. Both variants are well-established as associated with PD and have been reported in affected individuals. Of note, p.Asn409Ser is detected in the NBA, and carrier status of all sequenced carriers was concordant between the NBA and WGS data. No *GBA1* variant carriers exhibited co-occurrence with variants in other screened genes.

Most variants were extremely rare in gnomAD including the Latino/Admixed American subset, and had previous P/LP classification in ClinVar. However, one *GBA1* variant, c.1070C>T (p.Ala357Val), classified as LP, appears to be previously unreported, with no record in any of the assessed databases, including dbSNP, gnomAD, ClinVar, GP2, and GBA1 Browser. This variant was identified in a Colombian participant with an age at onset in their 30s.

Similarly, another LP *GBA1* variant, c.196G>C (p.Asp66His), identified in a Peruvian participant with EOPD and FHx+, including a deceased sibling affected by the disease, has not been reported in ClinVar or gnomAD. Interestingly, it has a previous record in dbSNP and was also identified in another affected participant from Peru within the GP2 WGS cohort.

Among variants located in PD genes associated with autosomal recessive phenotypes, six were observed in the compound heterozygous state (5 in *PRKN*, 1 in *VPS13C*), while one variant was observed in the homozygous state: *VPS13C* c.1340C>G (p.Ser447*) classified as LP. This variant has not been previously reported in the Latino/Admixed American subset of gnomAD or in ClinVar, though it was reported in two European (non-Finnish) carriers.

Only one synonymous variant was identified, *VPS13C* c.6480G>A (p.Lys2160Lys). Classified as LP, SpliceAI predicted a strong disruption of normal splicing, with high-confidence loss of both the donor (Δ score = 0.95) and acceptor (Δ score = 0.93) splice sites. This variant was identified in compound heterozygosity with *VPS13C* c.985C>T (p.Arg329*) in the same participant.

Among the genes associated with other forms of parkinsonism, all identified P/LP variants were observed in the heterozygous state. Interestingly, the *POLG* variant c.2246T>C (p.Phe749Ser) was identified in a participant who was also a carrier of a *PRKN* deletion of exon 4 (chr6:162111832-162217549).

A complete list of identified P/LP variants is presented in Table 2.

**Table 2.** Pathogenic and likely pathogenic variants meeting the expected inheritance pattern for the associated phenotype.

| Chr | Pos | Ref | Alt | rsID | Gene | Hets* | Homs | cDNA | Protein | CADD score | gnomAD | gnomAD AMR | GP2 AMR (PD) | ACMG Classification |
| --- | --- | --- | --- | --- | --- | --- | --- | --- | --- | --- | --- | --- | --- | --- |
| chr1 | 155235252 | A | G | rs421016 | *GBA1* | 5 |  | c.1448T>C | p.Leu483Pro | 25.3 | 0.00009858 | 0.00008342 | 0.005771567 | Likely pathogenic |
| chr1 | 155235727 | C | G | rs1064651 | *GBA1* | 1 |  | c.1342G>C | p.Asp448His | 24.1 | 0.0001033 | 0.0003005 | 0.0009113 | Pathogenic |
| chr1 | 155235843 | T | C | rs76763715 | *GBA1* | 5 |  | c.1226A>G | p.Asn409Ser | 23.8 | 0.001997 | 0.0009498 | 0.004252734 | Likely pathogenic |
| chr1 | 155239968 | GGTA | G | rs761621516 | *GBA1* | 1 |  | n.339+2_339+4 delTAC | p.Thr75del |  | 2.19E-05 | .00015 | - | Pathogenic |
| chr1 | 155239997 | C | G | rs1671993689 | *GBA1* | 1 |  | c.196G>C | p.Asp66His | 25.3 | 0 | 00 | 0.000303767 | Likely pathogenic |
| chr1 | 155236399 | G | A | - | *GBA1* | 1 |  | c.1070C>T | p.Ala357Val | 21.7 | 0 | 0 | - | Likely pathogenic |
| chr6 | 161785820 | G | A | rs34424986 | *PRKN* | 2 |  | c.823C>T | p.Arg275Trp | 26.1 | 0.0031 | 0.001849 | NA** | Pathogenic |
| chr6 | 161973418 | C | T | rs2485708259 | *PRKN* | 1 |  | c.619-1G>A | N/A | 33 | 7.03E-07 | 0.00001668 | NA** | Likely pathogenic |
| chr6 | 162443378 | CCT | C | rs55777503 | *PRKN* | 2 |  | c.101_102delA G | p.Gln34fs |  | 0.0003435 | 0.00006663 | NA** | Pathogenic |
| chr1<br>2 | 40310434 | C | G | rs33939927 | *LRRK2* | 1 |  | c.4321C>G | p.Arg1441Gly | 21.6 | 3.42E-06 | 0.00003342 | 0.001215067 | Pathogenic |
| chr1<br>5 | 61925456 | C | T | rs534039778 | *VPS13C* | 1 |  | c.6480G>A | p.Lys2160Lys |  | 1.44E-06 | 0.00007541 | - | Likely pathogenic |
| chr1<br>5 | 61991687 | G | C | rs2548060411 | *VPS13C* |  | 1 | c.1340C>G | p.Ser447* | 38 | 1.37E-06 | 0 | - | Likely pathogenic |
| chr1<br>5 | 62008659 | G | A | rs767007361 | *VPS13C* | 1 |  | c.985C>T | p.Arg329* | 36 | 5.55E-06 | 0.00006917 | - | Pathogenic |
| chr1<br>5 | 89322749 | G | A | rs769827124 | *POLG* | 1 |  | c.2419C>T | p.Arg807Cys | 32 | 2.74E-05 | 0 | 0.000303767 | Pathogenic |
| chr1<br>5 | 89323423 | A | G | rs202037973 | *POLG* | 1 |  | c.2246T>C | p.Phe749Ser | 27.3 | 2.67E-05 | 0.00003333 | 0.000303767 | Pathogenic |
| chr1<br>7 | 45983216 | C | T | rs146191766 | *MAPT* | 1 |  | c.637C>T | p.Arg213* | 22 | 3.99E-05 | 0.000152 | - | Likely pathogenic |
| chr1<br>7 | 46010418 | C | G | rs63751011 | *MAPT* | 1 |  | c.2091+16C>G | N/A |  | 0 | 0 | - | Likely pathogenic |

\*: heterozygotes for variants in genes with an autosomal recessive inheritance pattern are compound heterozygotes, Chr: Chromosome, Pos: Position, Ref: Reference Allele, Alt: Alternate Allele, rsID: Reference SNP cluster Identification, Het: heterozygote, Hom: Homozygote, cDNA: Coding DNA, CADD: Combined Annotation Dependent Depletion, AMR: Admixed American, ACMG: American College of Medical Genetics and Genomics
\*\* : Gene data not available in the GP2 Genome Browser

### Variants of uncertain significance

Eight VUS in *GBA1*, *LRRK2*, *PRKN*, and *VPS35* following appropriate MOI were identified in eight carriers (EOPD: 7, FHx+: 3, EOPD and FHx+: 4). All three *GBA1* variants (p.Ala495Pro, p.Gly103Ala, and n.-2158G>C) were observed in heterozygosity.

The single *LRRK2* VUS (p.Gly1312Glu) was also observed in heterozygosity. One participant carried two VUS *PRKN* variants (n.*514G>A and c.8-42026T>A) in compound heterozygosity, confirmed by statistical phasing. In *VPS35*, one participant was a heterozygous carrier of p.Val602Ile.

Eighteen VUS in the screened secondary parkinsonism genes were identified once each (EOPD: 8, FHx+: 5, EOPD and FHx+: 5), including one participant who carried both a homozygous variant in *PLA2G6* (p.Pro699Arg), as well as a homozygous intronic, non-coding variant in *MAPT* (n.-382G>A).

### *GBA1* risk variants

In *GBA1*, an additional two established risk variants, p.Thr408Met and p.Glu365Lys, were identified in three (EOPD: 1, FHx+: 1, EOPD and FHx+: 1) and four participants (FHx+: 1, EOPD and FHx+: 3), respectively. Both risk variants are detected in the NBA, and carrier status carriers were concordant between the NBA and WGS data. No *GBA1* variants exhibited co-occurrence with variants in other screened genes, although one carrier had both p.Glu365Lys and p.Thr408Met, as well as the P/LP p.Leu483Pro variant. The p.Thr408Met and p.Leu483Pro variants were on the same strand, with the p.Glu365Lys on the opposing strand.

A complete list of identified VUS and *GBA1* risk variants is presented in Supplementary Table 4. The distribution of P/LP variants and VUS across the primary PD-associated genes according to clinical subgroups is shown in Figure 2.

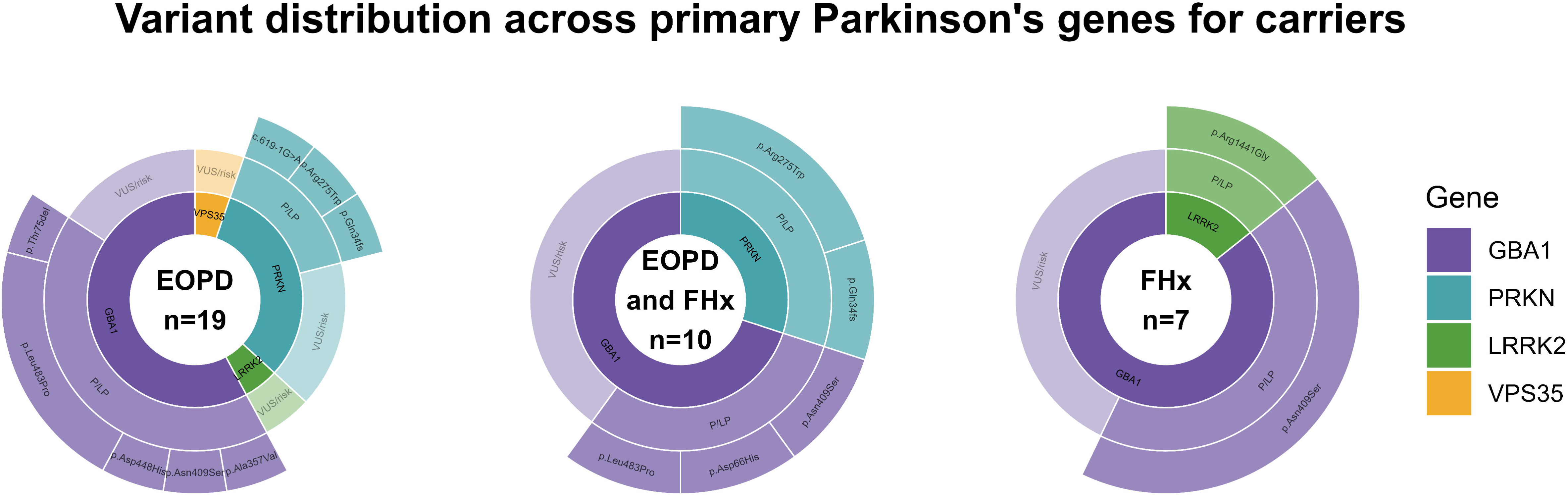

### Copy number variations

From the screened genes for CNVs, eleven unique CNVs were identified in *PRKN* (n=9) and *SNCA* (n=2) following appropriate MOI. All observed *PRKN* CNVs were deletions; one *SNCA* CNV was a duplication, and one was a triplication. CNVs in *PRKN* spanned between one and five total exons, while both *SNCA* CNVs spanned all exons. Four *PRKN* CNVs were observed in homozygosity, three in compound heterozygosity with a SNV in *PRKN*, and two with a second CNV in *PRKN*. Full specifics are presented in Supplementary Table 5.

### Presence of variants of interest in the GP2 Browser

The identified P/LP variants were cross-referenced using the GP2 Browser, paying particular attention to the Latino and Indigenous people of the Americas ancestry. Four *GBA1* variants– p.Leu483Pro, p.Asp448His, p.Asn409Ser, and p.Asp66His, as well as the *LRRK2* p.Arg1441Gly variant, were present in PD cases and absent in controls from Latin American populations. Both P/LP *POLG* variants, p.Arg807Cys and p.Phe749Ser, were also identified only in PD cases. The *VPS13C* and *MAPT* variants were absent from the Latin American dataset. The described *PRKN* variants were not reported in the GP2 Browser. Frequencies in Latin American populations in the GP2 WGS data are reported in Table 2 and Supplementary Table 3.

## DISCUSSION

In this study, short-read WGS identified P/LP in 12 of 256 (4.7%) Latin American PwP with EOPD and/or FHx+, and 14 of 256 (5.5%) with P/LP in *GBA1* for Gaucher’s disease who had previously tested negative using the NBA. Pathogenic and LP variants were identified in established PD-associated genes as well as in genes associated with other hereditary parkinsonian disorders. Moreover, several variants were previously unreported or absent from Latin American/Admixed American population databases, emphasizing the underrepresentation of these populations in current genomic resources and demonstrating the added value of WGS for improving the genetic characterization of PD in Latin America. In addition to P/LP variants, we identified several VUS that warrant follow-up.

Though our reported yield is lower than the monogenic burden reported by Mehanna et al. (∼20%), the numbers are not directly comparable, since our study participants were pre-filtered. Since some of the most common pathogenic variants, including *LRRK2* p.Gly2019Ser, were excluded from the sequenced cohort, we expect our yield to be lower than in cohorts that did not pre-filter pathogenic variants from genotyping data. From the EOPD and EOPD plus FHx+ combined, P/LP variants were identified in 4.7% of participants (EOPD: 4.4%; FHx+: 3.7%; EOPD and FHx+: 4.7%) and P/LP variants for Gaucher’s disease in *GBA1* were identified in 5.5% of participants (EOPD: 1.8%; FHx+: 0%; EOPD and FHx+: 1.6%) overall previously identified as P/LP negative in the NBA. The EOPD yield was much lower than previously reported in a predominately European cohort (13.8%) (31). The FHx+ yield was also lower than that reported previously in Latin America (12.7%) (9). However, neither of the results are directly comparable due to the pre-filtering in our study, as it is expected that our yield would be lower. This emphasizes that, while genotyping arrays can catch many common P/LP variants, especially those relevant in European populations, there is a monogenic burden missed, specifically in this Latin American population.

This also suggests that EOPD may represent a stronger clinical indicator of the presence of a disease-causing genetic variant than FHx alone, and that EOPD individuals may see more benefit from sequencing approaches. Additionally, since monogenic forms of EOPD are caused by rare variants, WGS provides greater sensitivity for detecting these variants than array-based genotyping. Together, these findings further add evidence to support current recommendations of prioritizing genetic testing in EOPD individuals; however, since this study did not include FHx-LOPD individuals, no conclusions can be drawn regarding the diagnostic yield of a similar approach in other groups of PD populations (3).

The highest burden was a result of variants in *GBA1* (P/LP: 14 carriers) and *PRKN* (P/LP: 5 carriers), similar to previous reports in Latin America, and *LRRK2* was relatively uncommon, likely due to exclusion of p.Gly2019Ser via the NBA (32).

Moreover, to date several variants seem to have been reported solely in Latin American individuals, suggesting possible population-specific or population-enriched variants. One example is the LP variant *PRKN* c.619-1G>A, identified in compound heterozygosity with a *PRKN* deletion in a Peruvian participant. This same variant was previously reported by Lorenzo-Betancor et al. in five Peruvian PwP, three of them also in compound heterozygosity and two homozygous (9). Furthermore, according to gnomAD, this variant has been reported only once in an individual from the Latin American/Admixed American subset. Although additional studies are needed to confirm its population distribution, the available evidence suggests that *PRKN* c.619-1G>A may represent a rare variant enriched in Latin American populations, specifically Peruvians. Expanding multi-center recruitment and detailed clinical phenotyping within local Andean populations will be critical to determine the true allele frequency and penetrance of this variant.

Similarly, the *GBA1* variant c.1070C>T (p.Ala357Val) appears to be previously unreported, with no records in the public databases assessed in this study. Interestingly, a different nucleotide substitution affecting the same codon, c.1070C>A (p.Ala357Asp), has previously been classified as likely pathogenic in ClinVar, suggesting that amino acid substitutions at this residue may be deleterious, aggregating evidence to the final classification of c.1070C>T as LP. The absence of p.Ala357Val from population databases, together with its identification in a Colombian participant, further highlights the value of WGS for identifying rare disease-associated variants in underrepresented populations, contributing to a better understanding of the genetic architecture of PD.

Several VUS identified in this cohort warrant additional discussion, despite not yet meeting requirements for their classification into another ACMG tier. In *GBA1*, the VUS p.Gly103Ala was notable since three other codon changes have been reported in the GBA1-PD Browser (p.Gly103Asp, p.Gly103Ser, and p.Gly103Val), though all reported variants are of unknown impact (GBA1-PD Browser). This recurrence may indicate a previously unrecognized mutational hotspot worth flagging for further functional studies. Additionally, in *GBA1*, p.Ala495Pro has been previously reported in MDSGene due to a functional study from 1994 (33); however, there are conflicting reports of pathogenicity in ClinVar. Similarly, the *VPS35* VUS p.Val602Ile is positioned only 18 amino acids from p.Asp620Asn, the only variant within the gene with established pathogenicity in PD. The VUS, with a CADD Phred score of 20.7, also falls within the region of the VPS35 protein that interacts with SLC11A2, the same region that the majority of possibly pathogenic *VPS35* variants fall within. Additionally, this variant was reported in a PwP in ClinVar, further supporting functional follow-up.

In the secondary parkinsonism-related genes, the homozygous variant in *PLA2G6* (p.Pro699Arg), observed alongside a homozygous variant in *MAPT* (p.Arg103Trp), appears to be novel in gnomAD and has a CADD score of 31, indicating it is predicted to be in the top 0.08% of most deleterious variants. The combination of homozygosity, absence in population databases, and high predicted deleteriousness warrants future functional follow-up.

Importantly, although both *POLG* variants identified in our cohort have previously been reported in individuals with recessive *POLG*-associated disorders who developed parkinsonism as part of their symptoms, they were observed in the heterozygous state in our participants (25203713). Consequently, their contribution to the PD phenotype cannot be established based on the available evidence. Whether heterozygous pathogenic *POLG* variants confer susceptibility to PD remains an area of ongoing investigation and warrants further study (42326775).

Finally, one VUS in *PRKN* (c.7+68242A>G) in an individual with an age at onset in their 30s was observed in compound heterozygosity with a known P/LP variant (p.Gln34fs). In autosomal recessive disorders, the identification of a VUS in trans with a known pathogenic variant represents additional evidence that may contribute to its future reclassification as P/LP if supported by complementary evidence, such as segregation studies, functional assays, or the identification of additional affected individuals carrying the same genotype. Therefore, continued evaluation of this variant as new evidence emerges may be warranted.

The challenges in accurately classifying some of the identified VUS may lie, at least in part, in the underrepresentation of Latin American populations in genomic studies. The limited inclusion of individuals with admixed Latin American ancestry has reduced the discovery and characterization of population-specific variants, thereby restricting the availability of the genetic, population, and clinical evidence required for accurate variant interpretation. Furthermore, the continued reliance on array-based genotyping approaches may limit the identification of previously unreported variants in underrepresented populations, further contributing to the challenges of variant classification. These findings highlight the value of implementing WGS to improve variant discovery and narrow this knowledge gap.

This study has several limitations. First, although this represents one of the largest WGS studies of monogenic PD in Latin American populations, the sample size remains modest and was restricted to eight countries, limiting the capacity to capture additional unreported variants and better characterizing the allele frequency of some others. Second, the pathogenicity of several of the newly identified variants was inferred mainly using ACMG/AMP criteria, public databases, published literature, and *in silico* predictions, requiring functional studies to completely characterize their pathogenic spectrum. Third, even though short-read WGS substantially improves variant detection when compared to array-based genotyping, it has limitations in detecting more complex structural variants, repeat expansions, and in complex regions of the genome, including those with pseudogenes, like *GBA1*.

In conclusion, these findings highlight the marked allelic heterogeneity of monogenic PD in Latin American populations and identify previously unreported or underrepresented variants that would likely be missed by array-based screening approaches. By expanding the spectrum of disease-associated variants identified in this population, this study contributes to a more comprehensive characterization of the genetic architecture of PD in Latin America and provides a valuable resource for future genetic and functional studies. Ultimately, these findings reinforce the imperative to transition from traditional array-based panels to comprehensive genomic sequencing in Latin American clinical practice, ensuring early and precise molecular diagnoses for patients with early-onset Parkinson’s disease.

## Data Statement

The data that support the findings of this study are available from the corresponding author, I.F.M., upon reasonable request. Data will also be available through GP2 in a future release.

## Acknowledgements

The authors would like to acknowledge all members of the LARGE-PD consortium, as well as, all participants, as well as their families, who contributed biospecimens and clinical questionnaires included in this work or support to the participants. The authors would like to acknowledge the Clinical Research Unit (UPC) of the Walter Cantídio University Hospital (HUWC), Federal University of Ceará, for its invaluable support in conducting this study. We are also grateful to the DNA-Neurogenetics Bank of the Instituto Nacional de Ciencias Neurológicas (BADN-Neurogenetics PERU) for supporting the collection of DNA samples and associated data used in this publication. We would also like to acknowledge the Center for Nanoscience and Nanotechnology, CEDENNA Project CIA250002. We would like to acknowledge Esther Colon, Dr. DaPrat G, and Espindola M for their hard work and dedication to LARGE-PD. Additionally, we would like to acknowledge the Molecular Biology Laboratory, Universidad Doctor Andrés Bello, San Salvador, El Salvador. This work made use of the High-Performance Computing Cluster provided by the Cleveland Clinic Research Computing Services HPC (and other Linux-based analytic resources, such as the large, stand-alone R, Python and GPU/CUDA servers) are supported by the Cleveland Clinic Research Computing Services. Our work would not have been possible without the support of Eldon Walker, Ph.D., Director, LRI Computing Services and Michael Weiner, Senior HPC Administrator.

## LARGE-PD consortium

Argentina: Emilia Mabel Gatto, Claudia Perandones, Martin Radrizani, Gustavo DaPrat, Natalia Gonzalez Rojas, Melisa Espindola, Martin Cesarini, Maria Valentina Muller, Carlos Matias López Razquin, Bibiana Pizarro, Lucia Wang, Clarisa Marchetti, Cesar Avila, Griselda Alvarado, Luciana Rojas-Vazquez, Juan Pablo Diaz-Rearte, Marcelo Kauffman, Sergio Rodriguez Quiroga, Dolores Gonzales, Pavel Alejandro Hernandez, Belen Ceballos, Florencia Echeverria, Gabriela Costa, Marcelo Merello, Federico Capparelli, Florencia Wainberg, Tomas Poklepovich, Florencia Mallou, Denise De Belder

Bolivia: Erick Gonzalez, Enrique Wagner, Robin Rodriguez, Alexander Quecana Janco Brazil: Bruno Lopes Santos-Lobato, Gracivane Lopes Eufraseo, Juliana dos Santos Duarte, Marcella Montenegro, Tatiane Souza, Camille Sena, Ândrea Ribeiro-dos-Santos, Pedro Braga-Neto, Deborah Rangel, Marconny Cavalcante, Mateus Balsells, Vitor Tumas, Angela Vieira Pìmentel, Gabriel R. Vilela, Joyce Yamamoto, Carolina P Souza, Ana L N Cunha, Vanderci Borges, Luiz Vinicius Silva Correa, Carolina Candeias da Silva, Henrique Ballali Ferraz, Dayany Leonel Boone, Mariana Cavalcanti Costa, Egberto Reis Barbosa, Grace Letro, Artur F S Schuh, Carlos R M Rieder, Gabriela Magalhães Pereira, Deise Cristine Friedrich, Thalya Osmilda Alves de Carvalho, Isabella Fonseca Benati, Jullivan Käfer Pasin, Vitor Picanço Lima Gomes, Marcelo Somma Tessari, Ingrid Lorena da Silva Gomes, Juan Sebastián Sánchez León, Paula Saffie-Awad, Gabriel Alves Marconi, Manoella Guatimuzim Testa da Silva, Eduardo Drews Amorim Colombia: Gonzalo Arboleda, Oscar Bernal Pacheco, Tatiana Lopez Gonzalez, Humberto Arboleda, Carlos Eduardo Arboleda Bustos, Hebert Bernal Castro, Juan David Caicedo Narvaez, Kelly Bonilla Vargas, Jorge Orozco, Beatriz Munoz Ospina, Harold Londono, David Aguillon, Sonia Moreno, Omar Buritica, David Pineda, Marlene Jimenez-Del-Rio, Carlos Vélez-Pardo, Sarita Firstman

Chile: Pedro Chana-Cuevas, Ximena Pizarro Correa, Consuelo Moos, Natalia Rojas, Patricio Olguin, Alicia Colombo, Juan Cristobal Nuñez, Andres De la Cerda, María Francisca Canals, Gonzalo Farías, Valentina Bessa, Mérida Terán, Pen Cheng Zhongxomg, Paula Saffie, Eduardo Perez, Dominga Berrios, Elías Fernandez, Marlene Valenzuela Valenzuela, Mario Fuentealba Sandoval, Susana Pineda, Floria Pancetti, Maria Eugenia Contreras Pinto, Benjamin Soto Flores

Costa Rica: Gabriel Torrealba-Acosta, Tanya Lobo-Prada, Jaime Fornaguera-Trías, Álvaro Hernandez-Guillen, Roger Rodríguez Dominican Republic: Rossy Cruz Vicioso, Ernestina Castro, Alpher Perez, Sergio Mosquera, Cesarina Torres, Janfreisy Carbonell Honduras: Reyna M. Durón, Alex Medina, Heike Hesse, Evelin Alvarez Herrera, Eduardo P. Murillo, Glenda Oliva

El Salvador: Susana Pena, Tatiana Ascencio, Oscar Peña Rodas, Ecuador: Faryd Llerena Toro, Michael Castelo, Carlos Rodriguez

Mexico: Mayela Rodríguez-Violante, Ana Jimena Hernández-Medrano, Amin Cervantes-Arriaga, Daniel Martinez Ramirez, Sarael Alcauter, Alejandra Medina-Rivera, Alejandra E. Ruiz-Contreras, Miguel E. Rentería, Alejandra Lázaro-Figueroa, Juan Manuel Esquivias-Farias, Andrés Morales-de-Arcia, Alejandra Zayas-Del Moral, Damaris Vazquez-Guevara, Yamil Matuk-Pérez, Carlos Manuel Guerra-Galicia, Ildefonso Rodriguez-Leyva, Karla Salinas-Barboza, Eugenia Morelos-Figaredo, Omar Cardenas-Saenz, Nadia A Gandarilla-Martínez, Sara Isais-Millán, Teresa Pérez-Torres, Domingo Martinez, Ingrid Estada-Bellmann, Roberto Trejo-Ayala, Carlos Alberto Ponce-Fernández, Dane Oropeza

Peru: Mario Cornejo-Olivas, Julia Rios Pinto, Angel Medina, Ivan Cornejo-Herrera, Koni Mejia-Rojas, Cintia Armas Puente, Edward Ochoa-Valle, Marcela Alvarado Morales, Elison Sarapura Castro, Andrea Rivera-Valdivia, Maryenela Illanes Manrique, Carla Manrique Enciso, Victoria Marca Ysabel, Olimpio Ortega Dávila, Freddy Requejo-Navarro, Alid Manrique Palomino, Gabriela Gushiken Oshiro, Laura Zelada Rios

Uruguay: Elena Dieguez, Victor Raggio, Andres Lescano

USA-Puerto Rico: Angel Vinuela, Esther Colon

USA: Thiago P Leal, Emily Waldo, Felipe Duarte-Zambrano, Miguel Inca-Martinez, Janvi Ramchandra, Kamilah Stark, Henry Mauricio Chaparro-Solano, Daniel Teixeira-dos-Santos, Emily Leininger, Nicolas Gutierrez, Valerie Rico, Anna E Anello, Emmanuel Scaria, Mariam Isayan, Paula Reyes-Pérez, Maria Rivera Paz, Ignacio F Mata, Karen Nuytemans, Anisley Martinez, Liena Infante

