## SupplementaryMethods for "Genomic Landscape of Early-Onset and Familial Latin American Parkinson’s Patients"

**Supplementary methods**

**Variant annotation**

Identified SNVs and indels in the genes of interest were annotated using the Ensembl Variant Effect Predictor (VEP) tool (1). Additional plug-ins to provide Combined Annotation Dependent Depletion (CADD) (2) scores and Genome Aggregation Database (gnomAD, version 4.1) (3) allele frequencies were utilized.

Moreover, annotations with the snpEff tool to retrieve functional consequences of the variants were performed. The first instances of functional characterization per variant were selected, including predicted impact severity and functional consequences. Additionally, we used the ANNOVAR (4) tool to assign rsIDs and retrieve variant pathogenicity classifications from ClinVar (version 20250721) (5); in silico prediction scores from dbNSFP (version 47 academic) (6) were also used. To characterize the deleteriousness of the variants, CADD score, MutationTaster (7), PrimateAI (8), AlphaMissense (9), SIFT (10), and PolyPhen2 (11) predictions were selected.

Lastly, to characterize the gene-disease relationships associated with the identified variants, the Online Mendelian Inheritance in Man database (OMIM) (12) was utilized to retrieve the Mendelian phenotypes and corresponding MIM numbers associated with each gene harboring a variant.
